# Risk factors for outcomes of COVID-19 patients: an observational study of 795 572 patients in Russia

**DOI:** 10.1101/2020.11.02.20224253

**Authors:** A.E. Demkina, S.P. Morozov, A.V. Vladzymyrskyy, V.G. Kljashtorny, O.I. Guseva, P.S. Pugachev, O.R. Artemova, R.V. Reshetnikov, V.A. Gombolevskiy, M.N. Ryabinina

## Abstract

**Background:** Several factors that could affect survival and clinical outcomes of COVID-19 patients require larger studies and closer attention.

**Objective:** To investigate the impact of factors including whether COVID-19 was clinically or laboratory-diagnosed, influenza vaccination, former or current tuberculosis, HIV, and other comorbidities on the hospitalized patients’ outcomes.

**Design:** Observational nationwide cohort study.

**Patients:** All subjects, regardless of age, admitted to 4,251 Russian hospitals indexed in the Federal Register of COVID-19 patients between March 26, 2020, and June 3, 2020. All included patients for which complete clinical data were available were divided into two cohorts, with laboratory- and clinically verified COVID-19.

**Measurements:** We analyzed patients’ age and sex, COVID-19 ICD-10 code, the length of the hospital stay, and whether they required ICU treatment or invasive mechanical ventilation. The other variables for analysis were: verified diagnosis of pulmonary disease, cardiovascular disease, diseases of the endocrine system, cancer/malignancy, HIV, tuberculosis, and the data on influenza vaccination in the previous six months.

**Results:** This study enrolled 705,572 COVID-19 patients aged from 0 to 121 years, 50.4% females. 164,195 patients were excluded due to no confirmed COVID-19 (n=143,357) or insufficient and invalid clinical data (n=20,831). 541,377 participants were included in the study, 413,950 (76.5%) of them had laboratory-verified COVID-19, and 127,427 patients (23.5%) with the clinical verification. Influenza vaccination reduced the risk of transfer to the ICU (OR 0.76), mechanical ventilation requirement (OR 0.74), and the risk of death (HR 0.77). TB increased the mortality risk (HR 1.74) but reduced the likelihood of transfer to the ICU (OR 0.27). HIV comorbidity significantly increased the risks of transfer to the ICU (OR 2.46) and death (HR 1.60). Patients with the clinically verified COVID-19 had a shorter duration of hospital stay (HR 1.45) but a higher risk of mortality (HR 1.08) and the likelihood of being ventilated (OR 1.36). According to the previously published data, age, male sex, endocrine disorders, and cardiovascular diseases increased the length of hospital stay, the risk of death, and transfer to the ICU.

**Limitations:** The study did not include a control group of subjects with no COVID-19. Because of that, some of the identified factors could not be specific for COVID-19.

**Conclusions:** Influenza vaccination could reduce the severity of the hospitalized patients’ clinical outcomes, including mortality, regardless of age, social, and economic group. The other factors considered in the study did not reduce the assessed risks, but we observed several non-trivial associations that may optimize the management of COVID-19 patients.

## 1 Introduction

According to the WHO monitoring, as of October 20, 2020, the COVID-19 pandemic counted 40,649,793 million confirmed cases globally, of which 1,126,538 were fatal [1]. Russia made a significant contribution to these numbers, with 1,415,316 total cases (4th place out of 217 indexed countries) and 24 433 deaths (13th place). At the time of writing, Russia’s incidence rate was 11 cases per 100,000 population [1], and hospital bed occupancy reached almost 90% [2].

Despite the high burden on hospitals, the national situation with new coronavirus infection is under control due to available reserve capacities [3]. There are no plans to reimpose rigid lockdown restrictions, and the Russian healthcare system continues to routinely perform testing, monitoring, and contact tracing of new cases. Federal recommendations for medical workers on the prevention, diagnosis, and treatment of COVID-19 have been developed to be followed by each hospital [4]. As our knowledge of the disease grows, causes of possible complications arise that could alter standard treatment guidelines.

Factors associated with the disease’s prognosis and outcome were the objects of several systematic reviews [5–7]. Unfortunately, most of the included studies have limitations associated with small or moderate sample sizes and single-center observations. There are very few publications that enroll large, nationwide, and unbiased cohorts of COVID-19 patients [8–10]. The published studies agree that the main risk factors associated with critical or mortal outcomes are demographic characteristics (age, sex, BMI) and comorbidities, such as hypertension, diabetes, cardiovascular disease, and respiratory diseases [7]. There are other not so well-studied but potentially important factors that require closer attention.

First, according to WHO guidelines, there are two different ICD-10 codes for the disease outbreak: U07.1 for COVID-19 confirmed by laboratory testing, and U07.2 assigned to a clinical or epidemiological diagnosis of COVID-19 [11]. To our knowledge, no comparison was made for clinical outcomes between the two groups of patients. Second, Marin-Hernandes et al. associated higher influenza vaccine uptake in adults aged 65+ years with lower COVID-19 mortality [12], which could also benefit other demographic groups. Third, there is no clear evidence on COVID-19 complications and mortality for the patients with tuberculosis (TB) [13, 14] or HIV [15]. The objective of this study was to investigate the factors including ICD-10 code, influenza vaccination, TB and HIV comorbidities that could impact on the length of a hospital stay, ICU and invasive mechanical ventilation requirements in relation with COVID-19 mortality on a nation-level sample of 541 377 subjects with completed inpatient treatment.

## Materials and Methods

### Study design and data sources

The study was approved by the Independent Ethics Committee of Moscow Regional Office of the Russian Society of Radiologists. The nationwide data from the Federal Register of COVID-19 patients (the COVID-19 Register) were used in this multicenter observational cohort study. The COVID-19 Register was established on March 31, 2020, by the Russian Government’s decision. The COVID-19 Register contains the data from all medical clinics and doctor’s offices of all constituent regions of the Russian Federation. The Register provides personified records on patients diagnosed with SARS-CoV-2 infection, subjects hospitalized with the signs of pneumonia, and those who have been in contact with these persons, including outcomes, form, duration, and extend of medical aid.

### Inclusion and exclusion criteria

All patients regardless of age, admitted to hospitals indexed in the COVID-19 Register were included in the present study. Diagnostic criteria were: (i) laboratory-confirmed SARS-CoV-2 infection by reverse transcription polymerase chain reaction (RT-PCR) detection of viral RNA (IDC code U07.1), or (ii) a clinical diagnosis based on the combination of signs of acute respiratory infection (ARI) and computed tomography (CT) results (IDC code U07.2). All included patients were divided into two groups: laboratory-verified COVID-19 (U07.1 cohort) and clinically verified COVID-19 (U07.2 cohort). Patients were excluded during data analysis if their clinical records did not contain sufficient data to access at least one of the following outcomes: overall survival, time to full recovery, need of ICU treatment or invasive mechanical ventilation (IMV) requirements.

### Variables Accessed

We analyzed patients’ demographic data (age and sex), ICD-10 code, the length of the hospital stay, and whether they required ICU treatment or IMV. The other variables for analysis were: verified diagnosis of pulmonary disease, cardiovascular disease, diseases of the endocrine system, cancer/malignancy, HIV, tuberculosis, and the data on influenza vaccination in the previous six months.

### Statistical analysis

We report means, standard deviation (SDs), medians, interquartile ranges (IQRs), minimum and maximum values for quantitative variables. The total number and proportion (%) of patients in each group were used for categorical variables. The aim of the analysis was to identify the relationship between the accessed variables and the outcomes using multivariable regression models. Two types of variables were used for the outcome assessment: (i) time-to-event variables for overall survival and time to discharge and (ii) binary variables for the need for ICU treatment and IMV. The first type of outcome variables was analyzed with a Cox proportional-hazards regression model with the inclusion of all the accessed variables as covariates. The risk ratio (Hazard Ratio, HR) of the event’s occurrence was calculated for each variable. All HRs were estimated with a 95% confidence interval (CI). For the second type of outcome variables, logistic regression models were created with an estimation of the odds ratio (Odds Ratio, OR) for each variable; ORs were also obtained with associated 95% CIs. P-values were determined by the Student’s t-test with significance threshold set at 0.05. We assessed time-to-event variables as time (in days) from hospital admission to either mortality or complete recovery. Patients who were still in hospital by the end of the observational period had their data censored at the latest follow-up date when it was evident that the event did not occur. Before the statistical analysis, we performed the dataset’s cleansing to remove entries with missing, invalid, and questionable data.

## Results

### Participants

We enrolled 705,572 COVID-19 patients aged from 0 to 121 years, 50.4% females, treated in 4,251 hospitals between March 26, 2020, and June 3, 2020 (Figure 1). 164,195 patients were excluded due to unconfirmed COVID-19 (n=143 357) or insufficient and invalid clinical data (n=20 831). In total, 541 377 participants were included in the study, 413,950 (76.5%) of them had laboratory-verified COVID-19, and 127 427 patients (23.5%) had clinical verification only (Table 1).

**Table 1.**
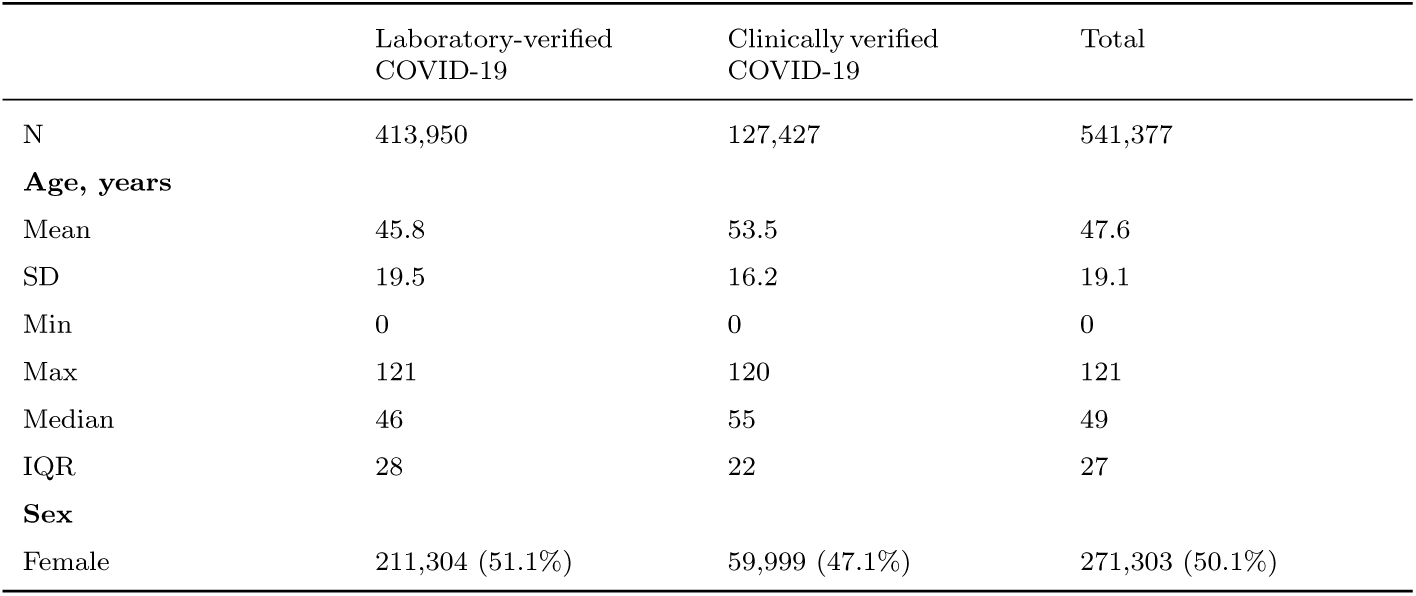
Demographics of the study participants

**Fig. 1.**
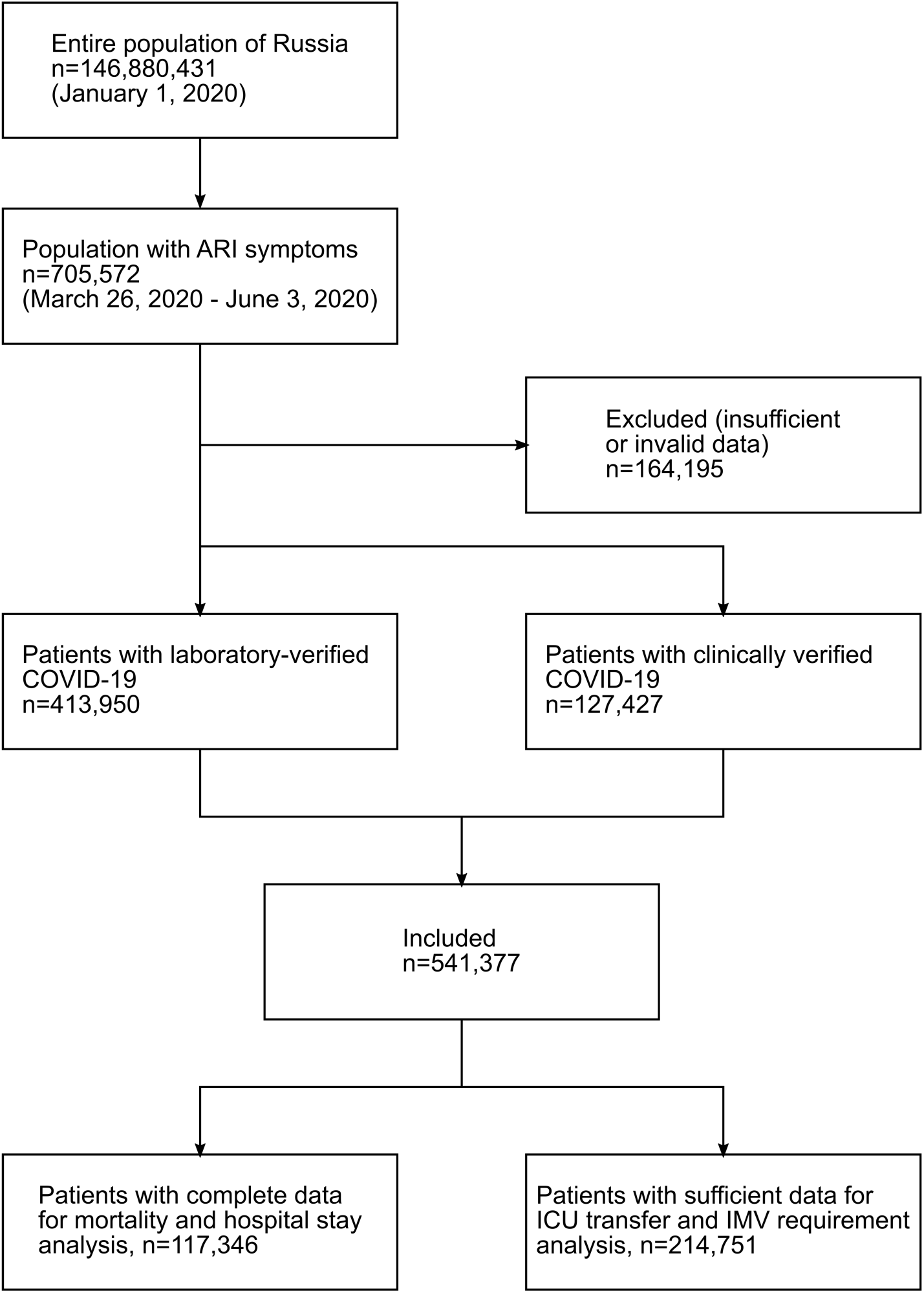
Flow diagram of participants through the study.

### Comorbidities

We observed comorbidities in 18.5% of the study participants: 17.5% with the laboratory-verified COVID-19 and 10.3% with the clinically verified disease.

The most common were cardiovascular diseases (8.2% and 10.3%; from now on, the first value refers to U07.1 patients and the second value corresponds to U07.2 patients), followed by diseases of the endocrine system (2.8% and 3.9%) and pulmonary diseases (1.7% and 2.1%). Among enrolled patients, 4.3% had several comorbidities (4.1% and 5.0%, Table 2).

**Table 2.** Comorbidities of patients with COVID-19

|  | Laboratory-verified COVID-19,<br>n=413,950 | Clinically verified<br>COVID-19, n=127,427 | Total, n=541,377 |
| --- | --- | --- | --- |
| **Comorbidity** |  |  |  |
| Pulmonary disease | 7,260 (1.75%) | 2,660 (2.09%) | 9,920 (1.83%) |
| Cardiovascular diseases | 33,966 (8.21%) | 13,194 (10.35%) | 47,160 (8.71%) |
| Endocrine system diseases | 11,737 (2.84%) | 5,000 (3.92%) | 16,737 (3.09%) |
| Cancer/malignancy | 3,830 (0.93%) | 943 (0.74%) | 4,773 (0.88%) |
| HIV | 330 (0.08%) | 145 (0.11%) | 475 (0.09%) |
| Tuberculosis | 273 (0.07%) | 51 (0.04%) | 324 (0.06%) |
| Other comorbidity | 14,982 (3.62%) | 5,578 (4.38%) | 20,560 (3.80%) |
| **Number of comorbidities** |  |  |  |
| At least one comorbidity | 57,716 (12.49%) | 19,874 (15.60%) | 71,590 (13.22%) |
| Two and more comorbidities | 16,798 (4.06%) | 6,350 (4.98%) | 23,148 (4.28%) |

### Length of the hospital stay

For the length of the hospital stay assessment, we analyzed the data of 117,346 discharged patients who had valid information on all factors included in the model. The length of the hospital stay was estimated as the number of days between the date of the patient’s inclusion in the COVID-19 Registry and their discharge or another outcome (death or transfer to another medical hospital). Patients still in hospital at the date of the data cutoff (June 3, 2020) were censored for the analysis. In total, the dataset contained 94,153 events of discharge and other outcomes. We performed the multivariate regression analysis using as covariates: patient’s gender, age, cohort (U07.1 or U07.2), the severity of COVID-19 expressed using the “CT 0-4” semi-quantitative grading scale [16], whether the patient received the ICU treatment or IMV, oxygen saturation level, comorbidities, and influenza vaccination status (Table 3). HIV comorbidity showed the least association with hospital discharge (HR 0.58, 95% CI 0.50 – 0.68, *p* < 0.001). Male sex was associated with a slightly longer length of the hospital stay (HR 0.96, 95% CI 0.95 – 0.97, *p* < 0.001). Age increased for every five years was also associated with a gradual increase in the length of the hospital stay (HR 0.98, 95% CI 0.97 – 0.98, *p* < 0.001). The factor most associated with the shorter length of the hospital stay was verified COVID-19 (HR 1.45, 95% CI 1.42 – 1.47, *p* < 0.001, compared to the laboratory-verified COVID-19).

**Table 3.** Length of the hospital stay, n=117,346

| Covariate | HR (95% CI) | *p-value* |
| --- | --- | --- |
| Age, with a five-year increment | 0.976 (0.974 - 0.978) | < 0.001 |
| Sex (ref: Female, HR 1) | 0.962 (0.950 - 0.975) | < 0.001 |
| Influenza vaccination | 1.022 (0.994 - 1.051) | 0.122 |
| **Primary diagnosis** |  |  |
| Clinically verified COVID-19 (ref: laboratory-verified COVID-19, HR 1) | 1.446 (1.422 - 1.470) | < 0.001 |
| **Comorbidity** |  |  |
| Pulmonary disease | 0.895 (0.865 - 0.925) | < 0.001 |
| Cardiovascular disease | 0.807 (0.791 - 0.823) | < 0.001 |
| Endocrine system disease | 0.879 (0.853 - 0.905) | < 0.001 |
| Cancer/metastasis | 0.884 (0.840 - 0.931) | < 0.001 |
| HIV | 0.582 (0.500 - 0.678) | < 0.001 |
| TB | 0.952 (0.797 - 1.137) | 0.587 |
| **Treatment** |  |  |
| ICU transfer | 0.409 (0.364 - 0.461) | < 0.001 |
| IMV | 0.119 (0.087 - 0.163) | < 0.001 |
| **COVID-19 signs** |  |  |
| An increase by one grade on the "CT 0-4" scale | 0.991 (0.975 - 1.008) | 0.307 |
| Oxygen saturation level, with 10% step | 1.070 (1.053 - 1.086) | < 0.001 |

### Mortality analysis

Among 117,346 participants with the full information on all factors included in the model, 4,996 patients had died at the data cutoff. Radiographic data were the most prominent indicators of the mortality risk. Disease progression by one gradation of the “CT 0-4” scale was associated with a five-fold increased risk of death (Table 4). Transfer to the ICU, history of TB, HIV, cancer, and male sex were also associated with mortality in descending order. The reduced risk of mortality was revealed in patients with co-existing pulmonary disease (HR 0.85, 95% CI 0.77 – 0.93, *p* = 0.001) and those who received IMV (HR 0.81, 95% CI 0.75 – 0.88, *p* < 0.001) and influenza vaccination (HR 0.77, 95% CI 0.63 – 0.95, *p* = 0.014). U07.2 patients had a slightly higher risk of death than U07.1 patients (HR 1.08, 95% CI 1.02 – 1.15, *p* = 0.014) (Table 4).

**Table 4.** Mortality risk factors, n=117,346

| Covariate | HR (95% CI) | *p-value* |
| --- | --- | --- |
| Age, with a five-year increment | 1.116 (1.104 - 1.129) | < 0.001 |
| Sex (ref: Female, HR 1) | 1.207 (1.140 - 1.277) | < 0.001 |
| Influenza vaccination | 0.775 (0.633 - 0.949) | 0.014 |
| **Primary diagnosis** |  |  |
| Clinically verified COVID-19 (ref: laboratory-verified COVID-19, HR 1) | 1.083 (1.016 - 1.154) | 0.014 |
| **Comorbidity** |  |  |
| Pulmonary disease | 0.847 (0.770 - 0.931) | 0.001 |
| Cardiovascular disease | 1.167 (1.093 - 1.246) | < 0.001 |
| Endocrine system disease | 1.174 (1.097- 1.257) | < 0.001 |
| Cancer/metastasis | 1.182 (1.065 - 1.311) | 0.002 |
| HIV | 1.599 (1.114 - 2.296) | 0.011 |
| TB | 1.737 (1.112 - 2.714) | 0.015 |
| **Treatment** |  |  |
| ICU transfer | 2.085 (1.892 - 2.297) | < 0.001 |
| IMV | 0.813 (0.752 - 0.881) | < 0.001 |
| **COVID-19 signs** |  |  |
| An increase by one grade on the “CT 0-4” scale | 5.280 (5.054 - 5.516) | < 0.001 |
| Oxygen saturation level, with 10% step | 0.935 (0.920 - 0.951) | < 0.001 |

### ICU transfer

The multifactor logistic regression model for analyzing ICU transfer data included fewer factors than the previous models. Therefore, the sample size of patients with valid clinical data counted 214,751 subjects, 8,018 of which required ICU treatment. The highest risk of the ICU transfer was associated with the oxygen saturation level (OR 43.26, 95% CI 40.38 – 46.34, *p* < 0.001) (Table 5). HIV, endocrine system diseases, pulmonary, cardiovascular diseases, and male sex were also associated with the risk of transfer to the ICU. Among the factors that could reduce the ICU requirement were influenza vaccination (OR 0.76, 95% CI 0.59 – 0.98, *p* = 0.031) and TB (OR 0.27, 95% CI 0.11 – 0.70, *p* = 0.007). There was almost no statistically significant difference between the U07.1 and U07.2 groups (OR 1.08, 95% CI 1.00 – 1.15, *p* = 0.046) (Table 5).

**Table 5.** ICU transfer risk factors, n=214,751*

| Covariate | OR (95% CI) | *p-value* |
| --- | --- | --- |
| Age, with a five-year increment | 1.051 (1.038 - 1.063) | < 0.001 |
| Sex (ref: Female, HR 1) | 1.162 (1.083 – 1.247) | < 0.001 |
| Influenza vaccination | 0.758 (0.589 – 0.976) | 0.031 |
| **Primary diagnosis** |  |  |
| Clinically verified COVID-19 (ref: laboratory-verified COVID-19, HR 1) | 1.076 (1.001 – 1.155) | 0.046 |
| **Comorbidity** |  |  |
| Pulmonary disease | 1.192 (1.026 – 1.386) | 0.022 |
| Cardiovascular disease | 1.181 (1.087 – 1.284) | < 0.001 |
| Endocrine system disease | 1.206 (1.082 - 1.344) | 0.001 |
| Cancer/metastasis | 0.871 (0.725-1.048) | 0.143 |
| HIV | 2.464 (1.380 – 4.399) | 0.002 |
| TB | 0.274 (0.107 – 0.703) | 0.007 |
| **Treatment** |  |  |
| ICU transfer | 1.289 (1.237 – 1.344) | < 0.001 |
| IMV | 0.813 (0.752 - 0.881) | < 0.001 |
| **COVID-19 signs** |  |  |
| Oxygen saturation level, with 10% step | 43.258 (40.381 - 46.339) | < 0.001 |

\*97,405 additional patients were included in this analysis due to the lower requirements for the completeness of clinical data

### Invasive mechanical ventilation

The multifactor logistic regression model was used to access the data of 214,751 patients, 4,736 of which received IMV. Similar to the transfer to the ICU, the oxygen saturation level was the leading risk factor (OR 26.58, 95% CI 24.81 – 28.49, *p* < 0.001) (Table 6). Endocrine system diseases and male sex also increased the risk of requiring IMV. The factors associated with reducing the risk of mechanical ventilation were cardiovascular diseases, cancer, and HIV (Table 6). Patients with clinically verified COVID-19 had a higher likelihood of being ventilated than the laboratory-verified cohort (OR 1.36, 95% CI 1.25 – 1.49, *p* < 0.001).

**Table 6.** IMV risk factors, n=214,751*

| Covariate | OR (95% CI) | *p-value* |
| --- | --- | --- |
| Age, with a five-year increment | 1.049 (1.034 - 1.064) | < 0.001 |
| Sex (ref: Female, HR 1) | 1.191 (1.093 - 1.297) | < 0.001 |
| Influenza vaccination | 0.742 (0.542 - 1.014) | 0.061 |
| **Primary diagnosis** |  |  |
| Clinically verified COVID-19 (ref: laboratory-verified COVID-19, HR 1) | 1.365 (1.251 - 1.489) | < 0.001 |
| **Comorbidity** |  |  |
| Pulmonary disease | 1.059 (0.890 - 1.260) | 0.517 |
| Cardiovascular disease | 0.840 (0.759 - 0.929) | 0.001 |
| Endocrine system disease | 1.295 (1.146 - 1.464) | < 0.001 |
| Cancer/metastasis | 0.760 (0.616 - 0.937) | 0.010 |
| HIV | 0.503 (0.247 - 1.023) | 0.058 |
| TB | 0.798 (0.328 - 1.939) | 0.618 |
| **Treatment** |  |  |
| IMV | 1.199 (1.155 - 1.246) | < 0.001 |
| **COVID-19 signs** |  |  |
| Oxygen saturation level, with 10% step | 26.585 (24.808 - 28.490) | < 0.001 |

\*97,405 additional patients were included in this analysis due to the lower requirements for the completeness of clinical data

## Discussion

Our hypothesis was that influenza vaccination, former or current TB, and HIV could be associated with the severity of COVID-19 outcomes. The results of our study revealed that influenza vaccination reduced the risk of ICU transfer (OR 0.76), IMV requirement (OR 0.74), and the risk of death (HR 0.77). According to Marin-Hernandes et al., one of the possible explanations of the negative correlation between influenza vaccination and COVID-19 mortality in the elderly Italian population could be a higher vaccine uptake in higher economic groups with better overall health [12]. We can confidently dismiss this statement due to our results based on vaccinated people of all age, social, and economic groups. The stimulation of innate immunity by an influenza vaccine could be the most feasible explanation of the observed correlation. Further research on the relationship between a recent vaccination and COVID-19 outcomes is needed to test this hypothesis.

TB increased the mortality risk (HR 1.74), but at the same time, the presence of TB reduced the likelihood of the ICU transfer (OR 0.27). Our findings contradict the statement made by Motta et al. that COVID-19 contributes to worsening the prognosis of TB patients and that TB might not be a major determinant of COVID-19 mortality [14]. The observed difference is possibly related to the different composition of the studied cohorts. In the study of Motta et al., there were only 6 cases from Russia in a cohort of 69 TB patients. Our sample included 324 TB patients. According to the estimates of Yusunbaeva et al., about 22% of patients with tuberculosis in Russia have multidrug-resistant strains [17]. In their study of 369 TB patients, the authors showed that the treatment failure could be as high as 82%, depending on the drug resistance category [17]. Therefore, our findings may be specific for the countries with a high proportion of drug-resistant *Mycobacterium tuberculosis* infections, such as Russia, South Africa, India, and the Philippines [18].

People living with HIV (PLWH) are at heightened risk for severe COVID-19 related complications compared to the general population [19]. While limited data are available on SARS-CoV-2 and HIV co-infection, chronic inflammation and immunodeficiency caused by HIV as well as HIV-associated comorbidities in older PLWH are associated with poor prognosis [20]. Our results have confirmed these concerns: HIV comorbidity significantly increased the risks of the ICU transfer (OR 2.46) and death (HR 1.60). The revealed fact that HIV infection was associated with a lower risk of IMV requires further investigations.

There was a statistically significant difference in outcomes between the patients classified with the U07.1 and U07.2 codes. The U07.2 patients had a shorter length of hospital stay (HR 1.45) but a higher risk of mortality (HR 1.08) and IMV treatment (OR 1.36). According to the Federal recommendations on the prevention, diagnosis, and treatment of COVID-19, a combination of ARI symptoms and CT findings was sufficient for the clinical diagnosis of COVID-19. The “CT0-4” semi-quantitative grading scale was used to evaluate the severity of the disease; immediate hospitalization was mandatory for the patients with CT3 and CT4 grades (pulmonary parenchymal involvement > 50%). Therefore, the U07.2 patients were admitted to hospitals at advanced stages of COVID-19-associated pneumonia. This fact could explain the shorter length of the hospital stay, IMV, and higher mortality.

Currently, one of the well-investigated risk factors for severe COVID-19 outcomes is the patient’s age. A study of 703 laboratory-confirmed COVID-19 patients admitted to 16 tertiary hospitals from 8 provinces in China reported that older age (> 60 years, HR 2.231, 95% CI 1.124-4.427, *p* = 0.022) was associated with increased odds of composite adverse outcomes such as in-hospital mortality, ICU admission, and IMV [21]. These findings were confirmed in a nationwide study of 1,591 COVID-19 patients admitted to 26 hospitals in Italy [22] and in the US study of 1 305 patients [23]. In some contradiction with that, Bello-Chavolla et al., in their study addressing 20 804 COVID-19 patients aged 60 years and older, demonstrated that a combination of age and comorbidities was a better predictor of COVID-19 severity and mortality than age alone [24]. Our study included patients of all age groups, and according to our findings, a five-year increment of the age of enrolled patients increased the length of the hospital stay (HR 0.98), the risk of death (HR 1.12), the risk of the ICU transfer (OR 1.05), and the risk of IMV treatment (OR 1.05).

Survival analysis revealed that male sex in patients with severe COVID-19 was associated with risk of hospitalization and ICU admission [24], severe illness (HR 1.87, 95%CI 1.43 - 2.46) or death (OR 1.97, 95% CI 1.29 - 2.99) [25–27]. Our findings are in good agreement with these data. Male sex was associated with the longer length of the hospital stay (HR 0.96), increased the risk of death (HR 1.12), ICU admission (OR 1.05), and IMV requirement (OR 1.05).

In accordance with the published data [5–7] our results show that endocrine disorders and cardiovascular diseases increased the length of the hospital stay, the risk of death, and the ICU transfer. Endocrine diseases were associated with a higher risk of IMV (OR 1.29) than cardiovascular pathologies, cancer or HIV (Table 6).

Our results revealed that cancer increases the length of the hospital stay (HR 0.88) and the risk of death (HR 1.18). The impact of concomitant oncological pathology depends on its severity, the presence of metastases, and therapy-induced complications. Despite that heterogeneity, our findings indicate that cancer is a significant risk factor that should be taken as a mandatory hospitalization criterion.

There was a contradiction regarding the influence of pulmonary diseases on the course of COVID-19. According to our results, pulmonary diseases’ presence reduced the risk of death (HR 0.85) and increased the risk of transfer to the ICU (OR 1.19). However, in previous studies conducted in Italy [28] and China [29], chronic obstructive pulmonary disease (COPD) was a significant predictor of mortality or factor of reaching the investigated outcomes. Contrary to that, Bello-Chavolla et al. showed that COPD was only associated with hospitalization [24]. One of our study’s limitations is that the COVID-19 Register does not contain the data on pulmonary disease codes. We believe that a detailed large-scale study is needed to address the effect of pulmonary disease’ type on the severity and outcomes of COVID-19.

The other limitation of our study is that the control group of subjects without COVID-19 was absent. Due to that, some of the factors that we have identified could not be specific for COVID-19.

Despite the limitations, our findings highlight several previously overlooked factors that could reduce the severity of outcomes and mortality and provide aid for the management of COVID-19 patients.

## Data Availability

The data used in this study cannot be made available in the manuscript, the supplemental files, or in a public repository due to Russian data protection laws.

## Acknowledgements

The authors express their gratitude to all doctors of medical organizations of the Moscow Department of Health, fighting the epidemic, and to the team of experts from the Moscow Department of Information Technologies for prompt assistance in working with data from UMIAS-ERIS.

